# Insights into motor impairment assessment using myographic signals with artificial intelligence: A scoping review

**DOI:** 10.1101/2025.04.02.25325099

**Authors:** Wonbum Sohn, M. Hongchul Sohn, Jongsang Son

## Abstract

Myographic signals can effectively detect and assess subtle changes in muscle function; however, their measurement and analysis are often limited in clinical settings compared to inertial measurement units (IMUs). Recently, the advent of artificial intelligence (AI) has made the analysis of complex myographic signals more feasible. This scoping review aims to examine the use of myographic signals in conjunction with AI for assessing motor impairments, while also highlighting potential limitations and future directions. We conducted a systematic search using specific keywords in the Scopus and PubMed databases. After a thorough screening process, 111 relevant studies were selected for review. These studies were organized based on target applications (measurement modality, modality location, and AI model purpose/application), sample demographics (age, sex, ethnicity, and pathology), and AI models (general approach and algorithm type). Among the various myographic measurement modalities, surface electromyography (sEMG) was the most commonly used. In terms of AI approaches, machine learning with feature engineering was the predominant method, with classification tasks being the most common application of AI. Our review also noted a significant bias in participant demographics, with a greater representation of males compared to females and healthy individuals compared to patient groups. Overall, our findings suggest that integrating myographic signals with AI has the potential to provide more objective and clinically relevant assessments of motor impairments.

## 1. Introduction

Motor impairments are prevalent in various clinical conditions such as stroke [1], spinal cord injury (SCI) [2], cerebral palsy (CP) [3], amyotrophic lateral sclerosis (ALS) [4], myopathy [5, 6], neuropathy [5, 6], multiple sclerosis (MS) [7], and Parkinson’s disease (PD) [8], significantly affecting patients’ ability to perform daily tasks and independence, and thus their quality of life. Objective assessment of motor impairments is crucial for enabling tailored care (diagnosis, treatment, and intervention) for such wide range of clinical population in need [9]. However, current clinical assessments often rely on subjective evaluations, leading to several limitations such as inter-rater variability [10] and ceiling effect [11]. On the other hand, laboratory-based quantification of motor impairment is not readily translatable to clinical settings, mainly due to practical barriers in transferring the technical resources (e.g., equipment, knowledge, and skills) required for acquisition, processing, and analysis/interpretation of the data collected [12]. There is an urgent need for objective clinical tools that can provide timely and precise assessments of motor impairments for effective intervention and precise medicine.

Recent advances in sensor and artificial intelligence (AI) technologies offer promising avenues for quantitative assessment of motor impairments in the real-world (e.g., clinical and/or daily setting) [13, 14]. As such, a large volume of studies in the past decade has focused on integrating AI with sensor-based measurements of human movement to recognize pattern/activity [15–23] or user intent [24–26], detect disease symptoms or adverse events [27–32], or provide bio-feedback during movement training/therapy [33]. Among the many sensor modalities used, inertial measurement units (IMUs) have dominantly been adopted, mainly owing to their compact size, low cost, ease of use (e.g. placement), and reliable performance [34, 35]. However, IMUs only measure motion, solely derived from kinematic parameters (i.e., translational acceleration, rotational speed, and orientation in space), and provide no information about the muscle activity or contraction that caused the movement, which often can result in little to no observable “motion” (i.e., isometric). For most, if not all, motor impairments, however, muscle activity is one of the most critical pieces of information for a comprehensive understanding of the underlying physiological mechanisms, as it is the ultimate manifestation of how the nervous system controls the physical part of the human body (e.g., limb segment).

We postulate that myographic signals — physiological activities measured from muscles — confer more than what IMU can offer, especially for motor impairment assessment. For example, myographic signals reveal complex physiological patterns such as coactivation [36, 37], fatigue [38–40], response to various types of sensorimotor stimulus [41, 42], motor unit recruitment [43], and the potential source (e.g., brain areas) governing the neural drive to the muscles [44, 45]. These insights cannot be captured through kinematic measurements alone and are crucial for understanding the pathophysiological mechanisms underlying abnormal movement patterns in clinical populations. By leveraging the information acquired with myographic signals, clinicians can detect subtle changes in muscle function and identify biomarkers that reflect the status of neuromuscular diseases, allowing for an evaluation of muscle function in real-world settings. Despite their significant potential, the technical challenges involved in acquiring, processing, and analyzing myographic signal (signal-to-noise ratio, location dependence, motion artifact, etc.) hinder the widespread adoption in clinical settings [46, 47]. Given the premise that AI is specialized in automated data processing/analysis and making prediction/inference based on potential patterns underlying large/complex set of signals, the AI-powered motor impairment assessment using myographic signals can address these limitations and offer a promising tool that provides more objective, precise, and clinically relevant information and insights for motor impairment assessment in clinical settings. Despite its promise, the use of myographic signals with AI models has received relatively little attention compared to IMU-based approaches, as evidenced by our preliminary literature search in Scopus and PubMed databases (Figure 1).

**Figure 1.**
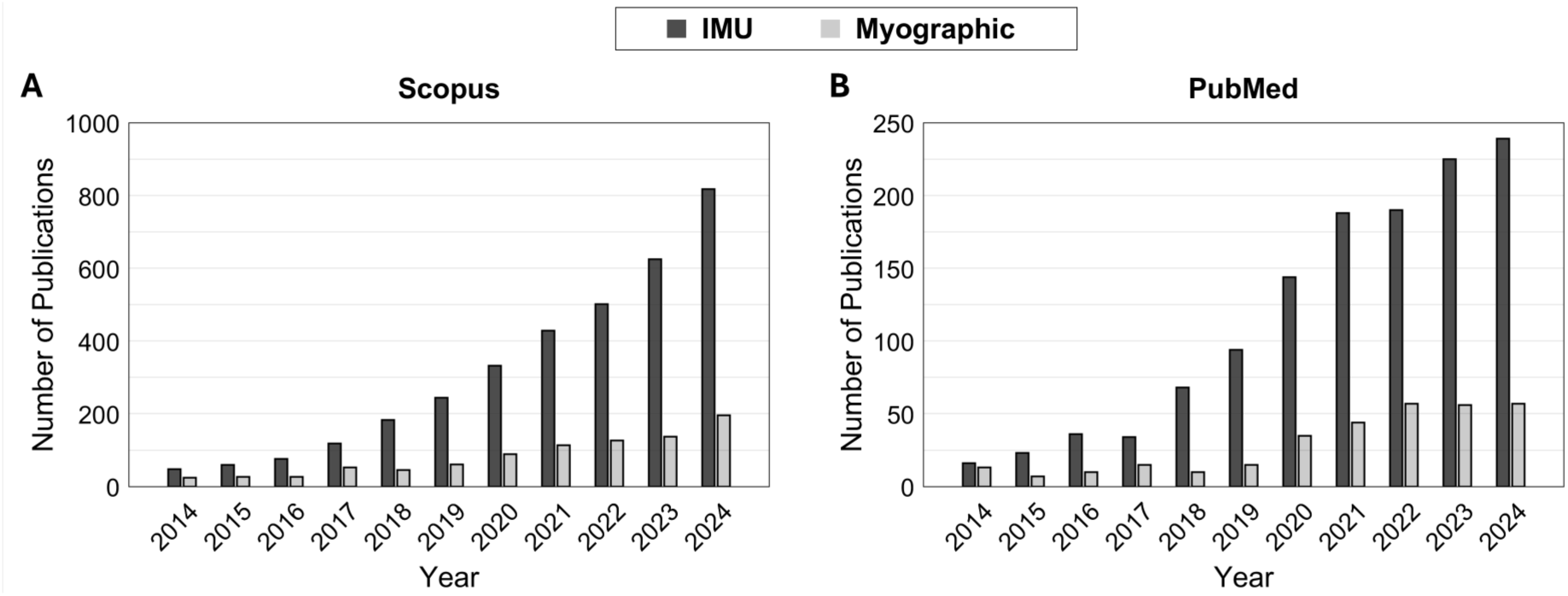
The number of publications from 2014 to 2024 in the Scopus (A) and PubMed (B) databases. The publications are categorized by measurement modality used: inertial measurement units (IMUs) vs. myographic signals. While there has been an evident rise in the number of research using each IMU and myographic signals, myographic signals have consistently received less attention than IMUs.

In the light of this knowledge gap, the purpose of this study is to provide a comprehensive overview of current use of myographic signals with AI for motor impairment assessment in the aspects of target application (e.g., classification, regression), sample demographics (e.g., age, sex, ethnicity, pathology), and AI model (e.g., machine learning, deep learning) and to discuss potential limitations and future directions for each of the above aspects.

## 2. Methods

A comprehensive literature search was conducted using Scopus and PubMed databases, followed by a study selection process in general accordance with the Preferred Reporting Items for Systematic Reviews and Meta-Analyses (PRISMA) guideline. The initial search was performed using combinations of the following terms: “AI”, “artificial intelligence”, “machine learning”, “deep learning”, “neural network”, “medical”, “clinical”, “patient”, “assessment”, “monitoring”, “diagnosis”, “tracking”, “impairment”, “motor”, “movement”, “force”, “torque”, “strength”, “kinematics”, “myogram”, “myography”, “EMG”, “MMG”, “FMG”, “OMG”, “SMG”, “ultrasound”, and “muscle”. The initial search results were further filtered with the following criteria: 1) restricting to works published within the last decade (2014–2024); 2) restricting to English-written, peer-reviewed journal article; and 3) excluding animal works. Finally, each study was manually screened, sequentially in the order by title, abstract, then full-text, based on the following criteria: 1) targeting clinical application; 2) measuring myographic signals; 3) utilizing machine learning and deep learning models; and 4) reporting results using myographic signals only. Additionally, relevant articles that satisfied the inclusion criteria were further identified through a manual search. The PRISMA flow diagram is shown in Figure 2.

**Figure 2.**
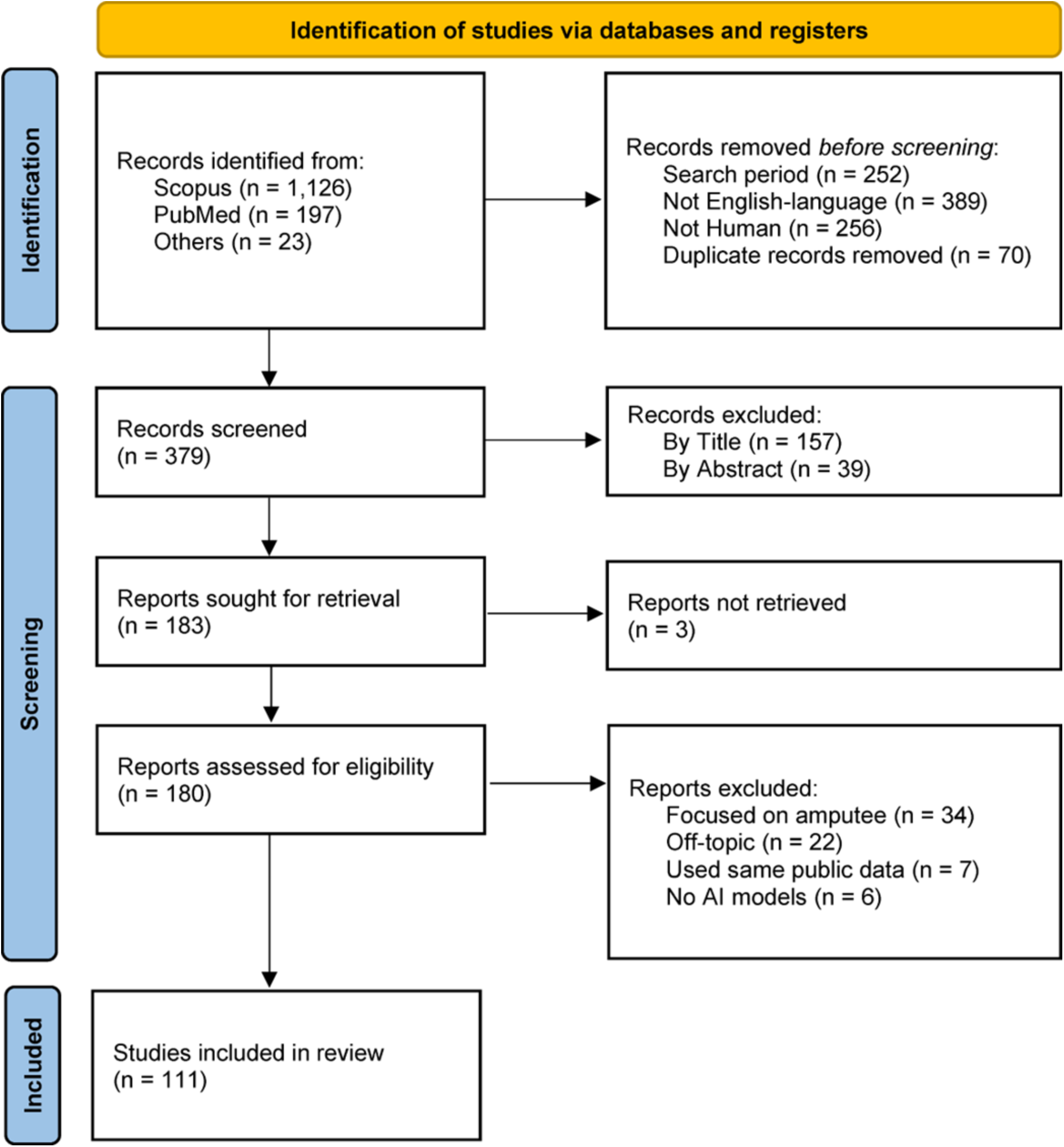
Overview of the PRISMA process for conducting a literature search, screening articles, and including relevant studies for this scoping review.

The selected studies were reviewed in detail, specifically focusing on the following scopes:

- *Target Application.* The selected studies were categorized into measurement modality, location (e.g., target muscle or joint), and purpose of the AI model/application. From detailed review of the selected studies with respect to this scope, we sought to determine whether the use of myographic signals indeed can provide more tailored insights into assessing motor impairment and/or allow for better performance of AI models compared to other measurement modalities such as IMU.
- *Sample Demographics*. Participant population in the selected studies were analyzed in terms of the portions between healthy individuals vs. patients, males vs. females, and ethnicities or origins of the populations based on study authors’ affiliations. In addition, we determined the age distribution among the healthy and patient groups in the studies where the mean and standard deviation values were reported, as well as the relationship between the number (i.e., absolute count) of male vs. female participants. From this scope, we sought to determine whether there exist any potential biases that may hinder the generalization of the developed/applied AI models, accounting for the heterogeneity in a particular clinical target population, or even broader populations in general.
- *AI Model*. The AI models used in the selected studies were categorized into general types of approach (i.e., machine learning with feature engineering, deep learning with feature engineering, deep learning without feature engineering) and specific algorithm. From this scope, we sought to determine whether the current approaches have potential implications for generalizability to diverse contexts of application.

In particular, we prioritized the studies that included patient populations for the above analyses in Target Application and AI Model.

## 3. Results

The initial literature search yielded a total of 1,346 studies. After restricting the search period from 2014 to 2024, 1,094 studies remained. Limiting the search to English-language articles further reduced the number to 705. Next, filtering for studies that focused on human subjects resulted in a selection of 449 studies. After removing duplicates, 379 studies were left. Based on the inclusion criteria, title screening reduced this number to 222, and abstract screening further narrowed it down to 183. Finally, after a full-text review, 111 studies were selected for the final analysis (Figure 1). Out of the selected 111 studies, 64 studies were conducted with only healthy participants [38–40, 46, 48–107], and 47 studies included data collected from patients [15–33, 108–135].

In the following sections, we provide a detailed review of the selected studies with respect to each of the three scopes and describe the results of the analysis proposed.

### 3.1 Target Application

Various measurement modalities were used to measure myographic signals (Figure 3), including surface electromyography (sEMG), intramuscular EMG (iEMG), high-density sEMG (HD-sEMG), sonomyography (SMG), mechanomyography (MMG), force myography (FMG), and optomyography (OMG). Among these modalities, EMG was the most frequently used measurement modality in all the reviewed studies (Figure 3A), which accounts for 79.5% including sEMG (67.2%) [15–20, 22–25, 28, 30, 38–40, 48, 50–68, 70, 72–74, 76–80, 82–89, 92, 93, 95, 96, 98, 99, 101–103, 105–122, 124–135], HD-sEMG (7.4%) [15, 17, 50, 67, 84, 86, 101, 120, 133], and iEMG (4.9%) [20, 27, 29, 31, 90, 123]. This is followed by SMG (8.2%) [21, 46, 57, 68, 79–81, 89, 91, 100], MMG (5.7%) [26, 32, 33, 55, 94, 104, 134], FMG (5.7%) [49, 52, 69, 71, 97, 107, 127], and OMG (0.8%) [75]. Most measurement modalities were also used in the studies including patient group (Figure 3B). There was a notable increase in the dominance of EMG by 7.5% — 2.8% from sEMG [15–19, 22–25, 28, 30, 108–122, 124–135], 0.6% from HD-sEMG [15, 17, 120, 133], and 5.1% from iEMG [20, 27, 29, 31, 123] — and of MMG by 2.3% [26, 32, 33, 134], while other measurement modalities including SMG (2.0%) [21], FMG (2.0%) [127], and OMG (0.0%) were less frequently used.

**Figure 3.**
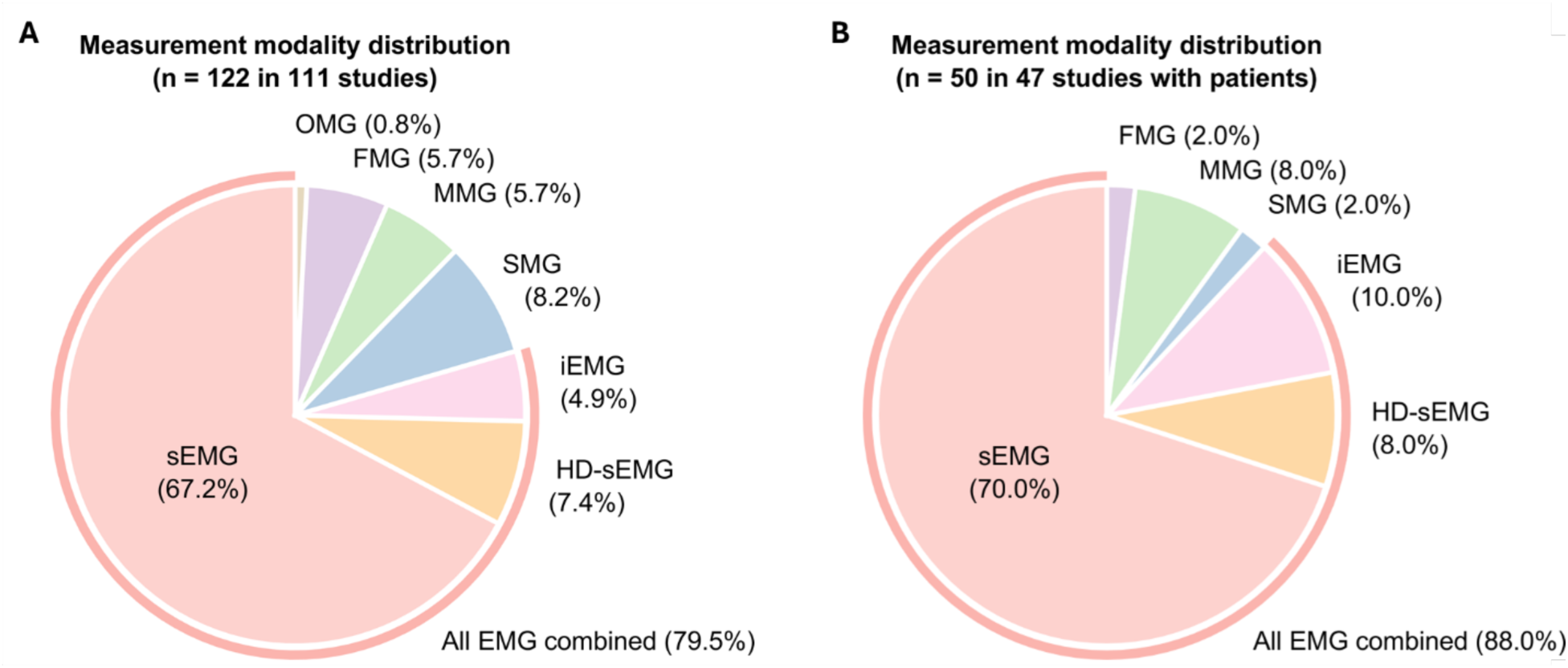
Distribution of measurement modalities used in the selected studies. (A) Among total 122 measurement modalities used across 111 studies, surface electromyography (sEMG) was the most prevalent modality, accounting for 67.2% of the total measurement modalities. Other modalities included sonomyography (SMG; 8.2%), high-density sEMG (HD-sEMG; 7.4%), mechanomyography (MMG; 5.7%), force myography (FMG; 5.7%), intramuscular EMG (iEMG; 4.9%), and optomyography (OMG; 0.8%). (B) Among total 50 measurement modalities used across 47 studies with patients, sEMG was the most dominant modality, accounting for 70.0%. Other modalities included iEMG (10.0%), HD-sEMG (8.0%), MMG (8.0%), SMG (2.0%), FMG (2.0%), and OMG (0.0%).

The patient-involved studies employed myographic signals with AI models mainly to perform classification and regression (i.e., prediction) applications, while the portion of such tasks varied by locations (Figure 4). Specifically, 78.4% of models in the studies focused on classification (Figure 4A) to identify various tasks including gestures [15, 16, 23, 112, 120, 122, 127, 133], movements or activities [17–19, 21, 22, 24, 110, 111, 116–118, 126], and clinical conditions such as diagnosis [27–32, 109, 115, 119, 123, 124, 131, 132] and severity [33, 125]. The regression task was also conducted in 21.6% of the studies to predict assessment score [111, 121], joint angle or torque [25, 26, 135], and muscle activation level or EMG values [129, 130]. In addition, these tasks were applied to various muscles located in peripheral upper and lower limbs as well as neck and torso (Figure 4B). Overall, lower limb was more frequently investigated compared to upper limb, and specifically at the ankle. Specifically, among the 47 studies that included patient populations, the tibialis anterior muscle was the most frequently assessed, appearing in 36.2% [19–21, 25, 29, 109, 114, 116–118, 123–125, 128–131], followed by the biceps brachii in 34.0% [17, 20, 21, 29–32, 108–111, 116, 121, 126, 128, 130], the rectus femoris in 21.3% [18, 29, 109, 114, 116, 117, 130, 131, 134, 135], the extensor digitorum in 17.0% [24, 112, 119, 121, 122, 127, 132, 136], the triceps brachii in 17.0% [17, 32, 108, 110, 111, 116, 121, 126], and the flexor carpi radialis in 17.0% [23, 24, 33, 110, 122, 126, 127, 132].

**Figure 4.**
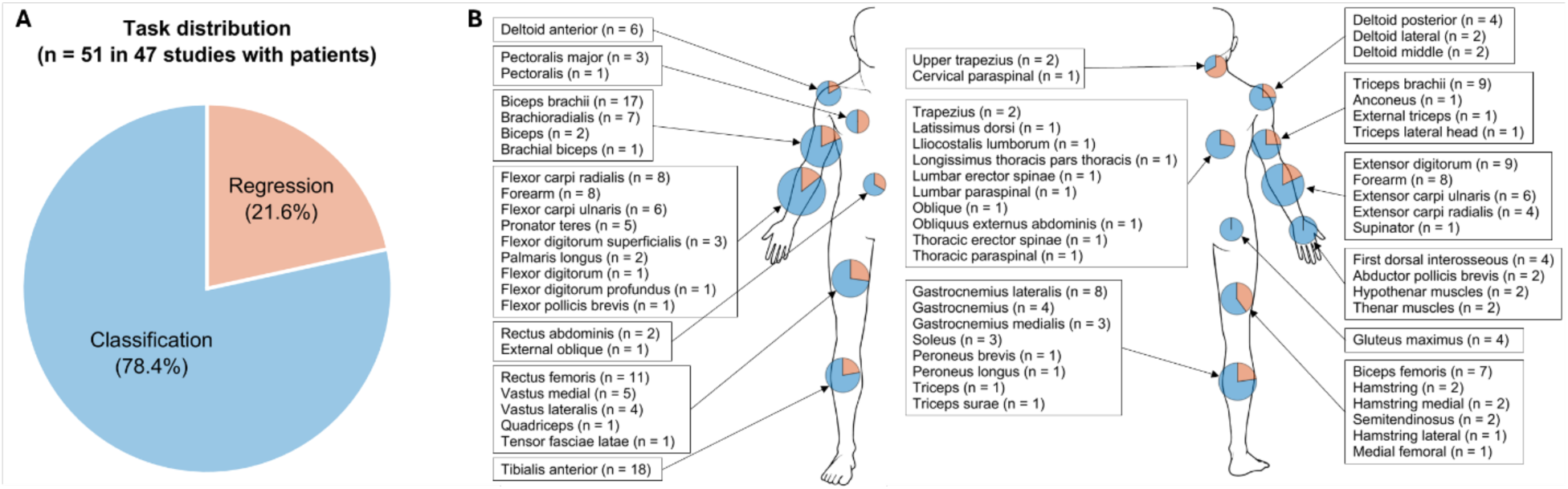
Applications and muscle group locations in the 47 studies involving patients. (A) Distribution or task. Among total 51 tasks reported, classification tasks accounted for 78.4% and regression tasks for 21.6%. Note that some studies addressed both classification and regression tasks. (B) Muscle group locations. Task proportions are depicted on a body schematic, with blue indicating classification tasks and red indicating regression tasks.

### 3.2 Sample Demographics

The total sample size was 2,541 in the selected 111 studies, and the number of patient participants was 933 (36.7%) from the 47 patient-involved studies [15–33, 108–135] (Figure 5A). Among the 47 studies that recruited patient participants, 29.8% aimed to balance the number of healthy and patient participants [21, 22, 25, 27, 28, 31, 108, 111, 114, 130–132, 134, 135]. Meanwhile, 34.0% focused exclusively on patient populations while applying various AI models with myographic signals [15, 17, 19, 20, 23, 24, 26, 33, 117, 118, 120, 121, 125–128].

**Figure 5.**
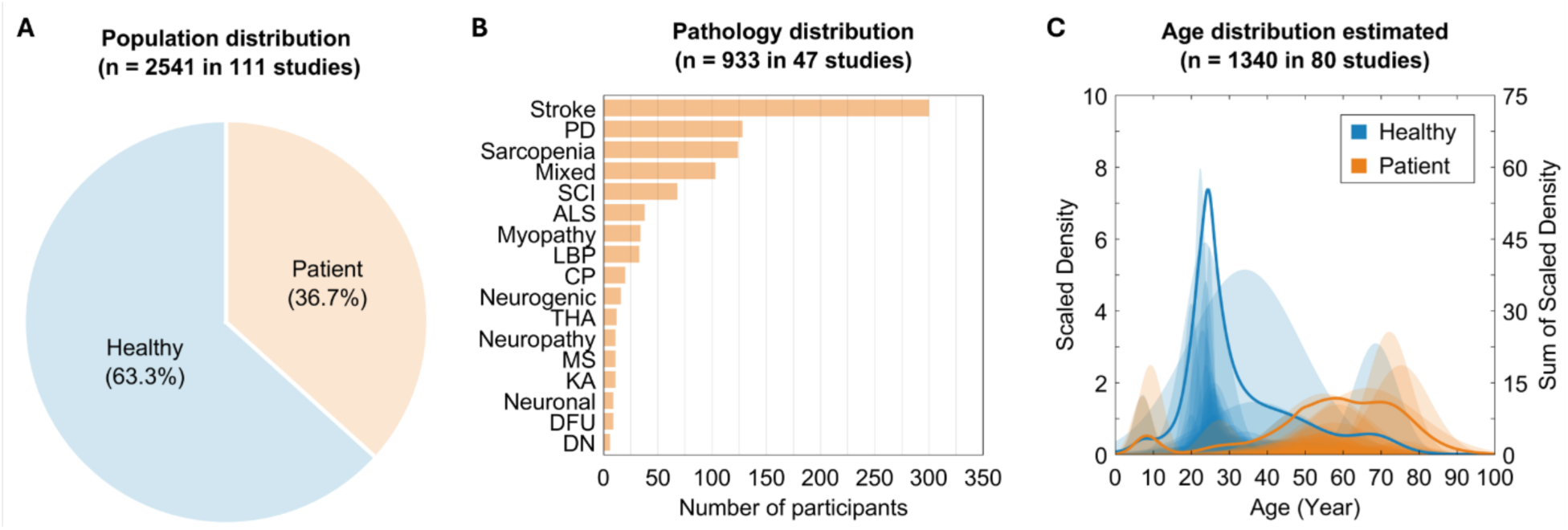
Demographic characteristics of study populations. (A) Distribution of healthy vs. patient population among total 2,541 participants across 111 studies. (B) Distribution of pathology among total 933 participants in the 47 studies with patients. PD: Parkinson’s disease; Mixed: radiculopathy, polymyositis, muscular dystrophy, peripheral nerve injuries, normal pressure hydrocephalus, and stroke; SCI: spinal cord injury; ALS: amyotrophic lateral sclerosis; LBP: low back pain; CP: cerebral palsy; THA: total hip arthroplasty; MS: multiple sclerosis; KA: knee abnormalities; DFU: diabetic neuropathy with ulceration; and DN: diabetic neuropathic. (C) Scaled density plot of the estimated age distribution across 80 studies with a total of 1,340 participants. The healthy group (blue) is predominantly younger, whereas the patient group (orange) exhibits a broader age range with a slight increase in density among older participants.

When looking at the pathology distribution (Figure 5B), stroke (32.2%; n = 300 in 16 studies [16, 18, 23, 24, 33, 108, 111, 113, 115, 116, 118, 121, 122, 126, 127, 129]) was the most dominant, followed by PD (13.7%; n = 128 in three studies [22, 32, 128]), sarcopenia (13.3%; n = 124 in three studies [125, 131, 132]), SCI (7.3%; n = 68 in ten studies [15, 17, 19, 25, 26, 110, 112, 120, 130, 133]), ALS (4.1%; n = 38 in four studies [21, 27, 119, 123]), myopathy (3.6%; n = 34 in six studies [27–31, 123]), low back pain (LBP; 3.5%; n = 33 in one study [109]), CP (2.1%; n = 20 in one study [117]), neurogenic (1.7%; n = 16 in two studies [29, 31]), total hip arthroplasty (THA; 1.3%; n = 12 in one study [114]), neuropathy (1.2%; n = 11 in one study [28]), MS (1.2%; n = 11 in one study [134]), knee abnormality (KA; 1.2%; n = 11 in one study [135]), neuronal (1.0%; n = 9 in one study [30]), diabetic foot ulcer (DFU; 1.0%; n = 9 in one study [124]), and diabetic nephropathy (DN; 0.6%; n = 6 in one study [124]).

Interestingly, the age distribution for healthy participants was highly focused on the young adults aged 20–30 years old (Figure 5C), while the age of patient participants was more broadly distributed as supported, in part, by the various target clinical populations (e.g., CP, SCI, stroke, neurodegenerative diseases), nevertheless, more focused on the middle age group (e.g., over 40 years old). Overall, discrepancy between the age distribution, both within and across studies, between healthy vs. patient population was evident.

The 81 studies that reported the number of male and female participants [15–17, 19, 21, 23, 27–33, 39, 40, 46, 48, 52–57, 59–65, 68, 69, 71–77, 79–91, 93, 94, 97, 100, 102–106, 108, 110, 111, 116–127, 130–132, 134, 135] recruited more male participants (n = 971; 58.6%) than female participants (n = 687; 41.4%) as a whole (Figure 6A). Specifically, 26.1% recruited a relatively balanced number of male and female participants, range of the female-to-male ratio of 40–60% [16, 17, 21, 27, 29, 33, 40, 46, 59, 62, 64, 69, 74, 75, 84, 87, 89, 97, 102–104, 106, 116, 117, 122, 123, 125, 131, 134]. There were also several studies with more female participants, range of the female-to-male ratio of 50–100% [15, 17–24, 26, 27, 29–33, 108, 109, 112, 114, 117, 118, 120, 121, 123–128, 130, 132]. Relationship between the number of male and female participants, however, revealed that in general even at an individual study level, the samples were biased towards more male participants (Figure 6B).

**Figure 6.**
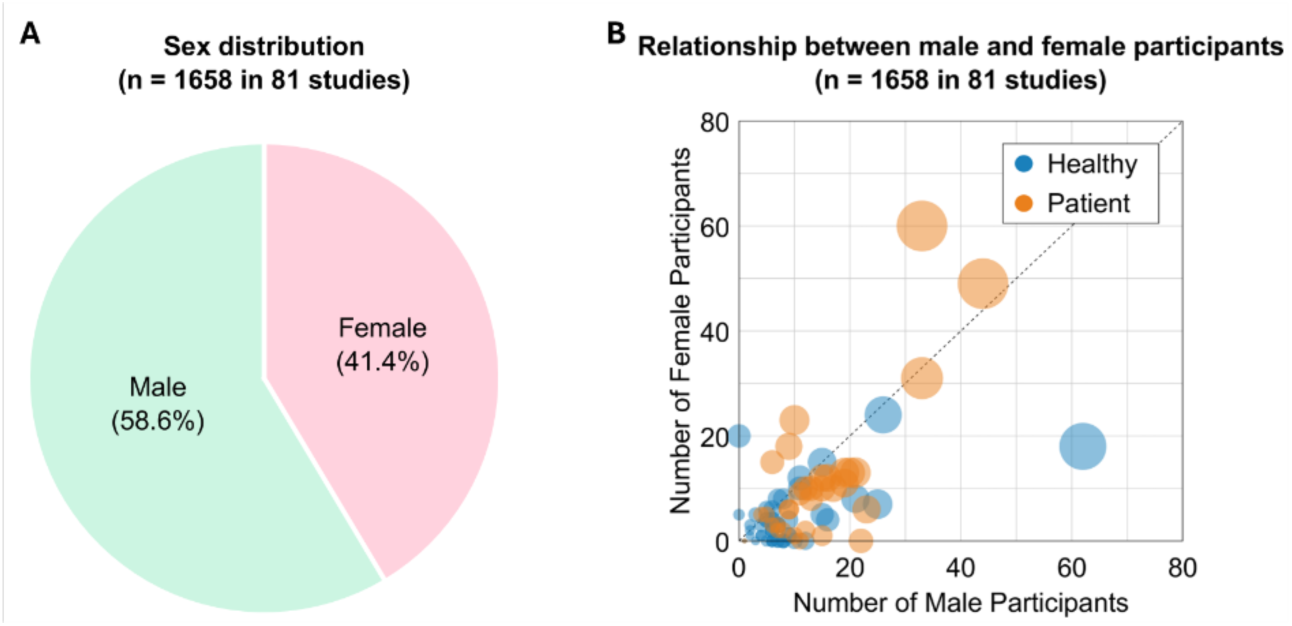
(A) Distribution of sex among total 1,658 participants in 81 studies where the number of both male and female participants was reported. (B) Relationship between male and female participants in the selected studies. Each circle represents an individual study, with circle size indicating total participant count. Blue and orange represent healthy and patient groups, respectively.

Affiliations of the authors in the selected studies — an estimate of ethnicity (or nationality) in the participant population — were highly biased to only few regions across the globe: mainly from northeastern Asia, western Europe, and north America (Figure 7A). Among the 227 different countries in the 111 studies (Figure 7B), 37.4% were from Asia [15, 18, 20, 21, 23–26, 28, 31, 38–40, 48, 50, 51, 53, 54, 56, 57, 59, 60, 63, 67, 70, 75, 77, 78, 82, 88, 90, 92–95, 101, 105–108, 111, 112, 115, 116, 118, 121–124, 126, 127, 129, 131, 132, 135], and 37.0% were from Europe [16, 17, 19, 23–25, 27, 29, 30, 32, 33, 50, 52, 55, 57–61, 63, 64, 68, 72, 73, 81, 83, 86, 87, 90, 96–99, 102, 103, 109, 114, 116–120, 125, 129, 133, 134]. North America accounted for 19.8% [15, 19, 22, 24, 46, 49, 66, 69–71, 73, 74, 76, 79, 80, 84–86, 89, 91, 93, 100, 104, 110, 112, 120, 128, 130], followed by Oceania for 3.5% [23, 26, 59, 62, 72, 86, 90, 118], South America for 1.3% [65, 82, 113], and Africa for 0.9% [97]. When considering the 47 patient-involved studies only (Figure 7C), Europe had the largest proportion at 43.4% [16, 17, 19, 23–25, 27, 29, 30, 32, 33, 109, 114, 116–120, 125, 129, 133, 134], followed by Asia at 40.4% [15, 18, 20, 21, 23–26, 28, 31, 108, 111, 112, 115, 116, 118, 121–124, 126, 127, 129, 131, 132, 135], North America at 12.1% [15, 19, 22, 24, 110, 112, 120, 128, 130], Oceania at 3.0% [23, 26, 118], South America at 1.0% [113], and Africa at 0.0%.

**Figure 7.**
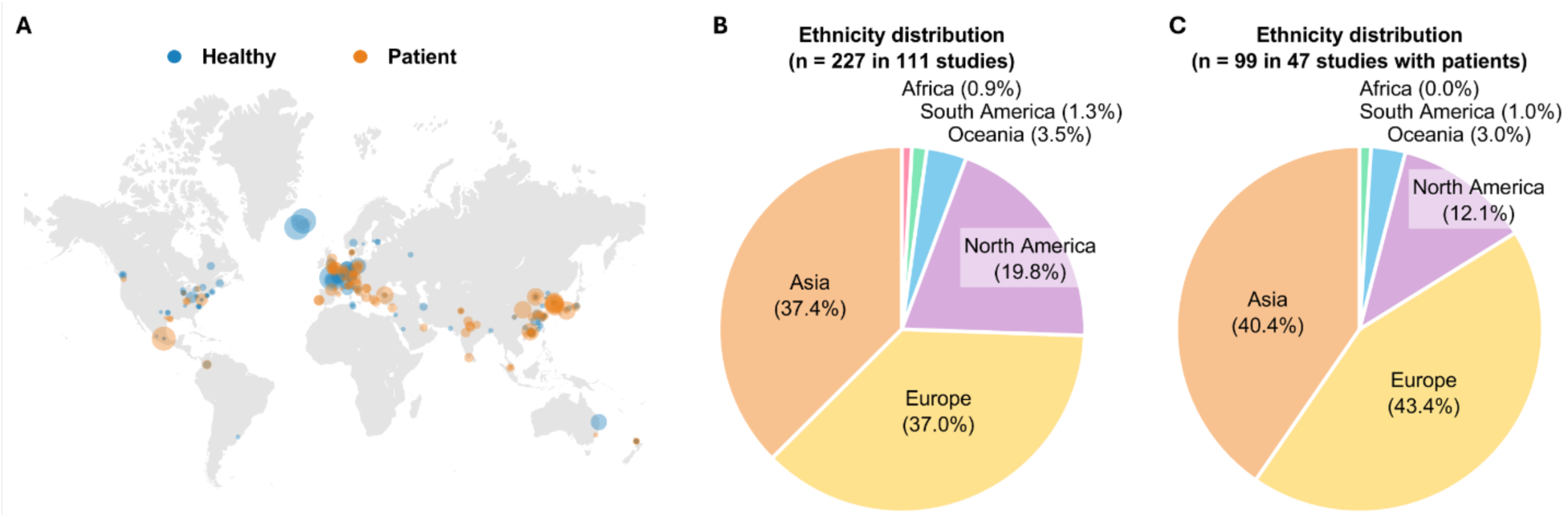
Distribution of geographic location and ethnicity based on the authors’ affiliations in the selected studies. (A) Global distribution, highlighting a higher concentration of the studies in North America, Europe, and Asia, with each circle representing an individual study and sized according to total participant count. Blue and orange represent healthy and patient groups, respectively. Distribution of ethnicity from 227 locations in 111 studies (B) and from 99 locations in 47 studies with patients (C). Note that multiple author affiliations per study result in a higher number of locations than studies.

### 3.3 AI Model

Various learning approaches and algorithms were used (Figure 8). Among the patient-involved studies, 90.3% employed various feature extraction and feature selection methods to utilize the measured myographic signals in different AI models [15, 17–20, 23–33, 108–111, 113, 115, 116, 118–127, 129–134]. Among the 28 studies utilizing neural networks (NNs), 64.3% further incorporated separate feature engineering techniques — while not necessary for deep learning models — for their analysis [24–28, 31, 32, 111, 113, 115, 118, 121, 123, 125, 126, 129–131]. In contrast, 35.7% used preprocessed myographic signals as direct input without additional feature engineering [16, 21–23, 112, 114, 117, 128, 133, 135].

**Figure 8.**
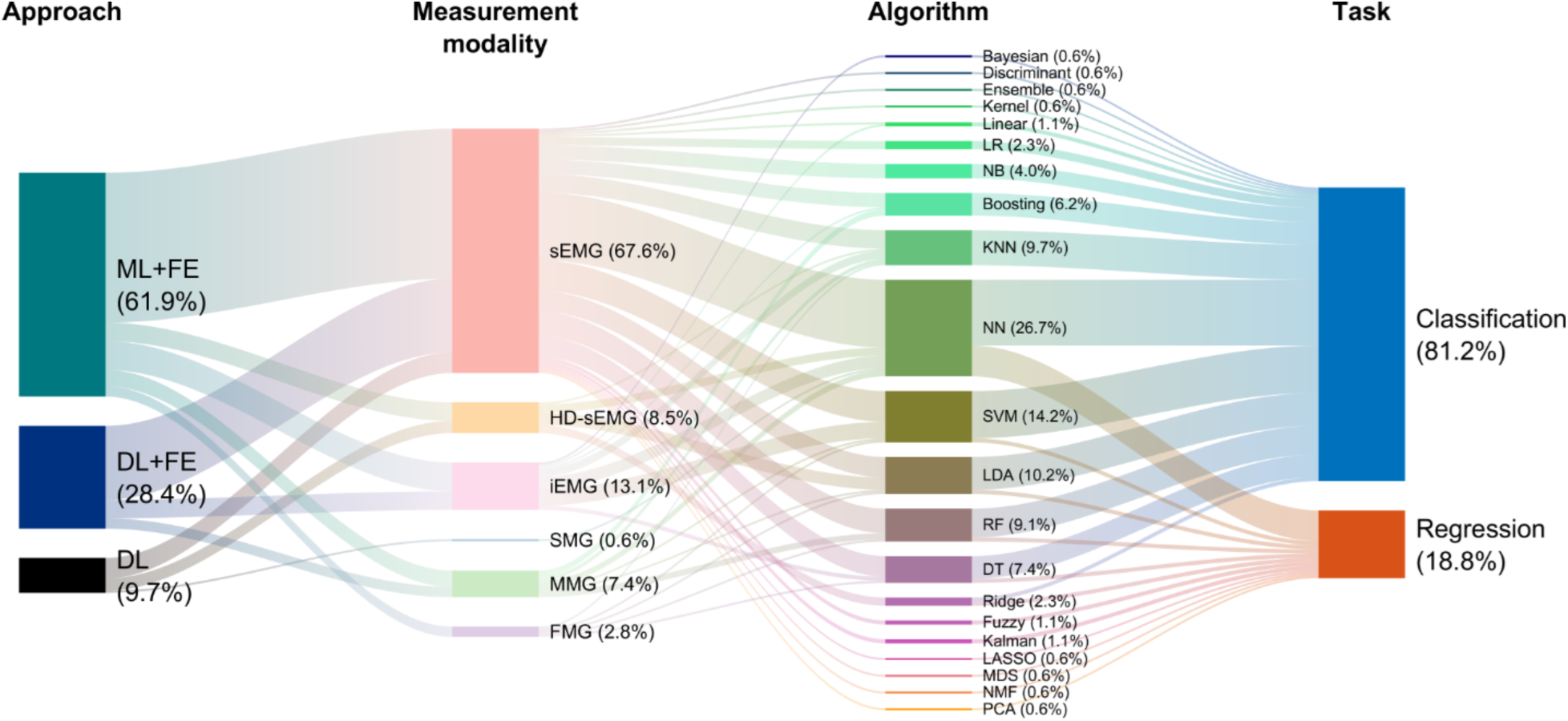
Sankey diagram illustrating the relationships between approach, measurement modality, algorithm, and application reported in the 47 studies with patients. This diagram visualizes the interconnections between different model approaches, including machine learning with feature engineering (ML+FE), deep learning with feature engineering (DL+FE), and deep learning without feature engineering (DL), and their applications in classification and regression tasks. sEMG: surface electromyography; HD-sEMG: high-density sEMG; iEMG: intramuscular EMG; SMG: sonomyography; MMG: mechanomyography; FMG: force myography; LR: linear regression; NB: naïve bayes; KNN: k-nearest neighbors; NN: neural network; SVM: support vector machine; LDA: linear discriminant analysis; RF: random forest; DT: decision tree; LASSO: least absolute shrinkage and selection operator; MDS: multidimensional scaling; NMF: non-negative matrix factorization; and PCA: principal component analysis.

In the 33 studies employing traditional machine learning methods for classification or regression tasks, 66.7% of the studies utilized sEMG [18, 19, 23, 25, 30, 108–111, 113, 115, 116, 118–120, 122, 124, 125, 127, 130–132, 134], 12.1% applied HD-sEMG [15, 17, 120, 133], and 15.2% incorporated iEMG [20, 27, 29, 31, 123] as their primary measurement modality.

Additionally, 9.1% investigated MMG [32, 33, 134], 3.0% focused on FMG [127], and 0% explored SMG. These studies mainly relied on conventional machine learning algorithms, such as k-nearest neighbors (KNN), linear discriminant analysis (LDA), support vector machine (SVM), decision tree (DT), or random forest (RF). In contrast, 28 studies that implemented deep learning techniques using NNs showed 75.0% focused on sEMG [16, 22–25, 28, 111–115, 117, 118, 121, 125, 126, 128–131, 135], 3.6% applied HD-sEMG [133], and 10.7% utilized iEMG [27, 31, 123].

Furthermore, 7.1% worked with MMG [26, 32], 3.6% with SMG [21], and 0% with FMG. Among the 39 papers related to classification, a survey of various AI models revealed that 69.2% employed NNs [15, 16, 19, 21–24, 27, 28, 30–32, 111, 112, 114, 115, 117–119, 123–128, 131, 133], 38.5% used SVM [18–20, 27, 29, 31, 111, 115, 116, 123–125, 127, 132, 134], and 25.6% utilized LDA [17, 23, 110, 118–120, 122, 127, 133, 134]. In contrast, among the 11 papers related to regression, 90.9% applied NNs [25, 26, 111, 113, 117, 121, 129, 130, 133, 135], 9.1% employed Support Vector Regression (SVR) [111], and 9.1% used Linear Discriminant Analysis (LDA) [133]. It is important to note that the total number and percentages are higher due to the use of multiple sensors or models within a single paper.

## 4. Discussion

In this review, we sought to provide evidence from current literature and state-of-the-art applications for whether using myographic signals with AI can offer better insights and performance in understanding and assessing motor impairments. Specifically, our goal was to conduct a scoping review of relevant works up to date using myographic signals with AI in terms of:

- *Target Application*, to determine whether the use of myographic signals indeed can provide more tailored insights into assessing motor impairment and/or allow for better performance of AI models compared to other measurement modalities such as IMU.
- *Sample Demographics*, to determine whether there exist any potential biases that may hinder the generalization of the developed/applied AI models, accounting for the heterogeneity in a particular clinical target population, or even broader populations in general.
- *AI Model*, to determine whether the type of the models being used have potential implications for generalizability to diverse contexts of application.

In the following sections, we review the main findings, discuss the limitations and remaining challenges, and propose potential solutions and future directions on the above aspects.

### 4.1 Target Application

In summary, we found that dominantly EMG, especially sEMG, is being used to acquire myographic signals (Figure 3), where the measurement took place in various body parts or muscles (Figure 4B), largely for classification tasks (Figure 4A).

In agreement with our postulation, we found promising evidence demonstrating that myographic signals can provide more direct insight into the muscle activity over other measurement modalities such as IMU that primarily capture movement-initiated patterns. Indeed, most of the reviewed studies demonstrated that the utilization of myographic signals with AI offers possibilities in: 1) the diagnosis of neuromuscular diseases, including myopathy [27–31, 123], neurogenic [29, 31], ALS [20, 21, 27, 119, 123], and DN [124] ; 2) the detection of abnormal muscle activity patterns from resting EMG signals [20] and from B-mode ultrasound images [91]; and 3) the assessment of the physical activity level in MS patients [134] and of the severity level of sarcopenia in older adults [125, 131, 132]. Additionally, we found some studies in the context of human-machine interfaces (HMIs), to detect/recognize user intentions [77, 137].

Interestingly, we also found studies that demonstrate the use of myographic signals in combination with other motion sensor modalities (e.g., kinematics, accelerometer, gyroscope, IMU) offering better performance, compared to just using myographic signals or motion signals, in classification [32, 111, 116, 128] and emulating clinical scores [111]. These findings suggest that different sensor modalities such as myographic and motion signals can complement each other for better (more accurate, robust) performance [32, 122].

Despite the promising evidence for the unique benefits that myographic signals can offer when used with AI for motor impairment assessment, there are remaining challenges. Owing to the inherent characteristics of EMG signals — capturing electrical action potential conducted through nerves — robust acquisition is relatively difficult, compared to other measurement modalities. In particular, non-stationarity of the signal, presence of noise from many sources [138, 139], as well as natural redundancy in how the same motor task can be performed using different motor commands [140, 141], induce variations (e.g., spatiotemporal, time and/or frequency domain) to the input for an AI model [142] and thus likely degrade the model performance (training vs. unseen dataset), especially for applications that are intended to be used for long period of time (e.g., across days or longitudinal) [143, 144]. In addition, due to the unique information based on frequency contents [145], acquisition requires relatively high sampling rate of ≥1 kHz (cf. usually ∼100 Hz for IMU), which demands for more power consumption and computational resources (e.g., processing, data storage). It is also worth noting that while other myographic signals based on SMG, MMG, FMG, and OMG — capturing physical changes in response to muscle excitation — can potentially overcome some of the limitations and may provide more robust inputs for AI models, these modalities also come with their own challenges, for example, related to signal-to-noise ratio and susceptibility to motion artifacts [146, 147].

Advances in sensor (e.g., materials, form factors, power/resource management) [148], signal processing techniques [145], and robust AI algorithm development [14] will be essential for overcoming these challenges. Another noteworthy future direction that this scoping shed light on is sensor fusion approach. Although the current review focused on the use of myographic signals in comparison to and implication with respect to motion data (e.g., IMU), other measurement modalities such as ECG, PPG, EDA, and/or EEG [149, 150] may provide additional, more comprehensive yet tailored insights into the physiological state underlying a specific motor impairment at an individual-specific, systemic level, complementing the unique motor perspective conferred by myographic signals; please refer to section 4.4. for discussions on the implications in terms of implementation.

### 4.2 Sample Demographics

In summary, significant demographic biases, particularly regarding age, sex, ethnicity, and pathology, were observed in the reviewed studies (Figures 5–7).

Given the established anatomical, biomechanical, and physiological differences across diverse populations, these biases would likely introduce variations in not only the input myographic signals that any AI model is being trained with but also the (often latent) features being captured/learned. Consequently, such biases may limit the generalizability of the developed application to broader target and ultimately hinder the clinical translation. Implication of such differences across diverse populations is increasingly gaining attention in science, probably due to heterogeneity across individuals. For example, there are measurable sex-/ethnicity-based differences in inherent neuromuscular performance such as body composition (e.g., muscle mass and fat distribution) [151–153], muscle strength and power [152, 154, 155], muscle architecture (e.g., fascicle length, pennation angle, muscle thickness) [156, 157], and muscle fiber characteristics (e.g., fiber type, cross-section area) [158, 159]. Furthermore, age-related changes, exercise adaptations, and pathological conditions can lead to even greater diversity in neuromuscular mechanisms including motor unit firing behaviors (e.g., firing rate, recruitment) [160–162], muscle fiber conduction velocity [163–165], muscular changes in size (e.g., atrophy, hypertrophy) [158, 166, 167], architecture [168–170], material properties (e.g., composition of adipose tissue and fibrous collagen in extracellular matrix) [171–173], and fiber type composition [158, 164, 174], and muscle coordination [175–177]. In addition, lifestyle-related factors such as physical activity, nutrition, and comorbidities may further introduce the variability of myographic signals [178–180].

The inclusion of diverse populations is essential to enhance the generalizability of research findings across a wide range of individuals and contexts. However, it is well-acknowledged that acquisition of such a comprehensive dataset practically is nearly impossible for any individual investigator or research lab, especially for clinical population [181]. Such challenge can be overcome with effort as a community, such as openly sharing data (e.g., repository, database), which, encouragingly, seems to be the recent trend in many disciplines [182–184]. In order to maximize the potential of such combined effort, standardized protocols for data acquisition and processing are essential [182]. Moreover, the integration of advanced techniques, such as data augmentation leveraging generative AI models [185, 186], may provide valuable insights. Nevertheless, it is essential to carefully consider the methodological implications and caveats associated with these approaches, including potential biases and limitations in data quality, validity, and reliability. Additionally, while longitudinal, real-world tracking of quantitative motor impairment-related data, including myographic signals, is becoming more accessible with the advances in wearable sensor and remote monitoring techniques [187, 188], a care must be taken in protecting healthy-related and privacy information [184].

### 4.3 AI Model

In summary, we found that machine learning with feature engineering is the most dominant category of AI models that are being used with myographic signals for clinical applications (Figure 8). It was interesting to note that deep learning models, which by virtue does not necessarily require a priori definition of specific features to learn from the input dataset [189], were more often used with feature engineering. We also found that neural network is the most widely used model type/architecture, where, in many cases, various models were used in one study to compare the performances.

The performance of an AI model trained with relatively small data (e.g., sample size) with respective to model complexity (e.g., number of features or parameters) as well as for particular purpose (e.g., classification or prediction) will likely not generalized to other data set or application [190]. While feature engineering can improve the performance of AI models, it may potentially increase the risk of overfitting [191]. In addition to ensuring the diversity in the input data/sample discussed above (in section 4.2), there are approaches that can be adopted to improve the generalizability and robustness of the AI model for broader contexts of application. For example, transfer learning is a scheme that leverages cross-domain techniques to generalize a model pre-trained with initial source data/domain to newly recorded target data/domain without the necessity for complete retraining or recalibration of the model [192]. Successful examples, in the context of hand gesture classification, include retaining accuracy across EMG data measured from different users, sensor locations, and days (within the same user) [193]. Alternatively, various model-specific/agnostic explainable AI techniques and tools applied at local/global scope (e.g., SHapley Additive exPlanations (SHAP) or Local Interpretable Model-Agnostic Explanations (LIME) [194]), may allow for identification of key features that can be adapted to guide and facilitate the generalization of a particular AI model to a different set of data or application (e.g., patient, clinical target). Compared to other disciplines and applications, such approaches have been rarely applied for AI models using myographic signals, especially for clinical target [195, 196].

While our initial intent was to also investigate, among the studies reviewed, the effect of model complexity, such as by examining its correlation with the sample size and/or performance, we could not find a single, suitable measure for model complexity that can be commonly applied to all models reviewed [197]. Moreover, many studies did not report the basic information about the AI model (e.g., architecture, size) from which we can infer the complexity [24, 27, 126]. At the minimum, if not tested explicitly, it is encouraged that such information is provided to aid in gauging the generalizability of the model. Furthermore, we assert that the development of universal/versatile measures and means to evaluate the model complexity is needed, which, analogous to the established power analysis tools for statistics, can inform and ultimately guide the selection of type, size, structure/architecture of AI models to use.

### 4.4 Clinical translation

Ultimately, we emphasize the following two important aspects to be considered, and implemented, for any application using myographic signals with AI to find its place in the real world, that is, deployed in the field (e.g., clinics, bedsides, home) and adopted by the users (e.g., clinicians, patients, and their caregivers). Firstly, the technology as the entire package should be user-friendly. For example, the sensor/device should be easy to “do-on-and-off” (i.e., easily/quickly placed without much care), requiring minimal (ideally single) placement and setup. In case of multi-modal measurements or sensor fusion, recent advancements in sensor integration technology appear to be promising to pack multiple sensors onto a single, smaller chip [149]. The control/software interface should also be simple and intuitive, requiring minimal to no technical knowledge and/or training for clinicians and patients to easily use [198]. Secondly, the model outcomes should provide clinically relevant information. Whether providing a very close link (e.g., strong correlation) to the conventional clinical assessment measures or newly devised outcome metrics, the information gathered/synthesized must readily translate to what clinicians currently use and correspond to what the patient experiences in everyday life [199, 200].

## 5. Conclusion

In conclusion, this scoping review highlights the promising application of myographic signals with AI in understanding and assessing motor impairments. Through an extensive search of the Scopus and PubMed databases, our analysis demonstrated that sEMG is the predominant measurement modality for acquiring myographic signals, mainly used for classification tasks, and that machine learning with feature engineering is the most common AI approach employed in clinical applications, including identification of neuromuscular diseases. Moreover, our findings showed significant demographic biases within and across studies, suggesting the need for more diverse and representative datasets. We also discussed two important aspects to translate this effort of using myographic signals with AI into real-world clinical practice. Ultimately, we believe that myographic signals, given the essential physiological information it conveys at high spatial and temporal resolution, combined with AI approaches that robust and accurate performance offers great potential for precision medicine in the context of motor impairment assessment.

## Data Availability

All data produced in the present study are available upon reasonable request to the authors.

